# Questioning the axiom: Is hyperflexed scaphoid a common occurrence in South Indian population? - An observational study

**DOI:** 10.1101/2022.05.14.22275085

**Authors:** Mohamed Nazir Ashik, Maithreyi Sethu, Srinivasan Rajappa

**Author notes:** Corresponding Author Name : Dr.Maithreyi SETHU, Permanent Address : No 19, 6^th^ Cross street, Lake area, Nungambakkam, Chennai, Tamil Nadu, India 600034, Present Address : No 19, 6^th^ Cross street, Lake area, Nungambakkam, Chennai, Tamil Nadu, India 600034, E-mail Address, Fax Number.

## Abstract

**Background:** Scapholunate angle SLA greater than 80° is one of the indicators of surgery for carpal instability. The normal SLA is between 30-60°. This is based on studies which were conducted several decades ago in the western part of the world. The SLA of Indian population has never been measured before. The knowledge of normal SLA in each population becomes imperative because it lays down the guidelines for surgical and non-surgical management of wrist fractures and instability in that population. In this study, we aim to measure the SLAs in asymptomatic volunteers of South Indian origin and compare the results with existing data.

**Methods:** An observational study was done in a tertiary care hospital in South India. Lateral radiographs of the wrist of asymptomatic volunteers were taken. The SLA was measured using a software in the PACS system. The relation of SLA to gender, age, left and right sides and handedness was compared. These were compared to pre-existing global data available on SLAs.

**Results:** The SLA of the 202 radiographs with 47 males and 54 females were studied.

- The average SLA was 51.33° and the range was between 29° and 74°.
- The average SLA of males was 51.8° and females 50.8°. There was no significant difference between the two.
- Similarly, there was no significant difference between the SLA of right and left sides in right handed and left handed subgroups.
- However, there was an increasing trend in SLA among females with age. But no similar pattern was seen among males.
- Interestingly, 39 out of the 202 radiographs (19.3%) had a SLA above 60°.

**Conclusion:** This study suggests the normal SLA in South Indians may be higher than the previously established reference values and serves as a pilot project and a reference for further studies on SLA.

## INTRODUCTION

Anthropometry is the measurement of living human individuals to understand the range of normal human physical variation.^5^ To achieve accurate sex and age determination of normal values, every population should have its specific measurements.^6^ The normal range of the wrist measurements vary with race and ethnicity. The scapholunate angle plays an important role in the decision making of management of certain carpal bone and scapholunate ligament injuries. With peri-lunate, wrist and hand injuries accounting for nearly 28% of all injuries^7^, it is interesting to know what is the normal range of the SLA of different ethnicity.

The present study is the first to measure the scapholunate angles (SLA) of healthy South Indian population. The normal range of scapholunate angle (SLA) is between 30-60^o 123^. This is based on studies which were conducted several decades ago in the western part of the world. The SLA of Indian population has never been measured before. The knowledge of normal SLA in each population becomes imperative because it lays down the guidelines for surgical and non-surgical management of wrist fractures and instability in that population. In this study, we aim to measure the SLAs in asymptomatic volunteers of South Indian origin and compare the results with existing data.

## METHODS

A descriptive observational study was conducted in a tertiary care centre in South India between January 2021 and July 2021. The aim was to measure the SLA in a normal, asymptomatic South Indian population and analyse any correlation with age, gender, between right and the left sides and handedness. Statistical analysis using average, standard deviation and range, significance of correlation of SLA with the different variables were calculated using the null hypothesis. Institutional ethical clearance was obtained before starting the study (CSP-MED /21/SEP/71/129). Inclusion criteria were, patients above 20 years but less than 60 years with no specific complaints in upper limb, who were clinically normal and healthy volunteers. Excluded were children in whom epiphysis would not have closed, patients with acute or chronic wrist pain, recent or previous injury to the wrist, hand or forearm. Congenital conditions affecting the hand or wrist bones, infections and tumours of the hand and those with associated systemic condition causing hypercalcaemia or hypocalcaemia were also excluded from the study. Standard lateral radiographs of both the wrists were taken.

The scapholunate angle was measured using the software in PACS. These lateral radiographs were taken in standard ***Zero position*** with the arm adducted against the trunk, the elbow flexed 90^0^ with the forearm in neutral rotation (no supination or pronation), and the wrist in neutral position (no radial or ulnar deviation, and no flexion or extension).^9^ We attained this using a below elbow thermoplastic splint as shown in the diagram (Fig 1). It is similar to the Meyrueis’ technique where a wooden board was used to maintain the forearm in a strict lateral position.^10^ Rotational malalignment alters the carpal bone measurements^11^ and by using this easy technique we attained the neutral wrist position. Scapholunate angle was measured using the software in our PACS (Fig 2). The

**Fig 1:**
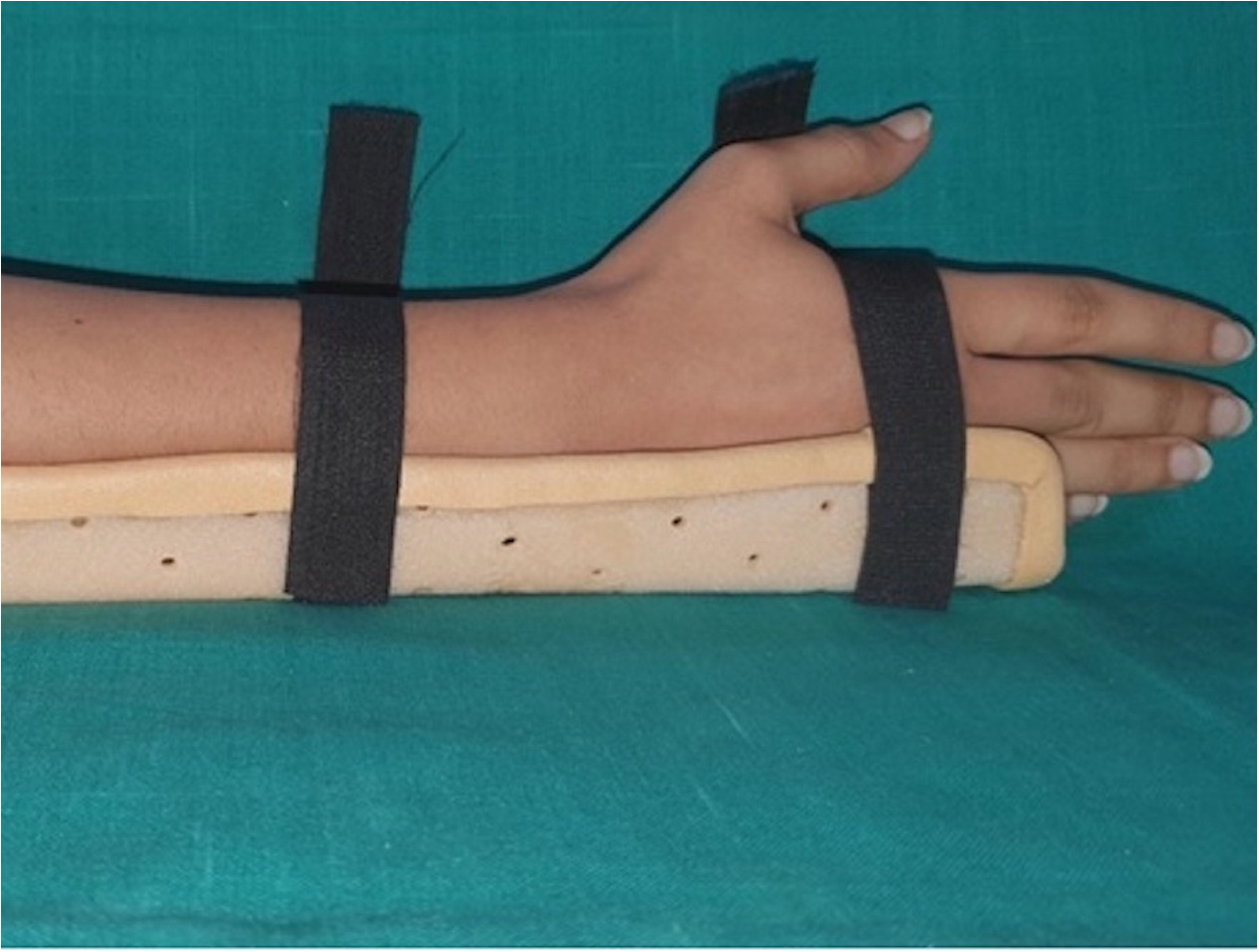
Thermoplastic splint aiding in keeping the wrist in neutral position during the radiograph

**Fig 2:**
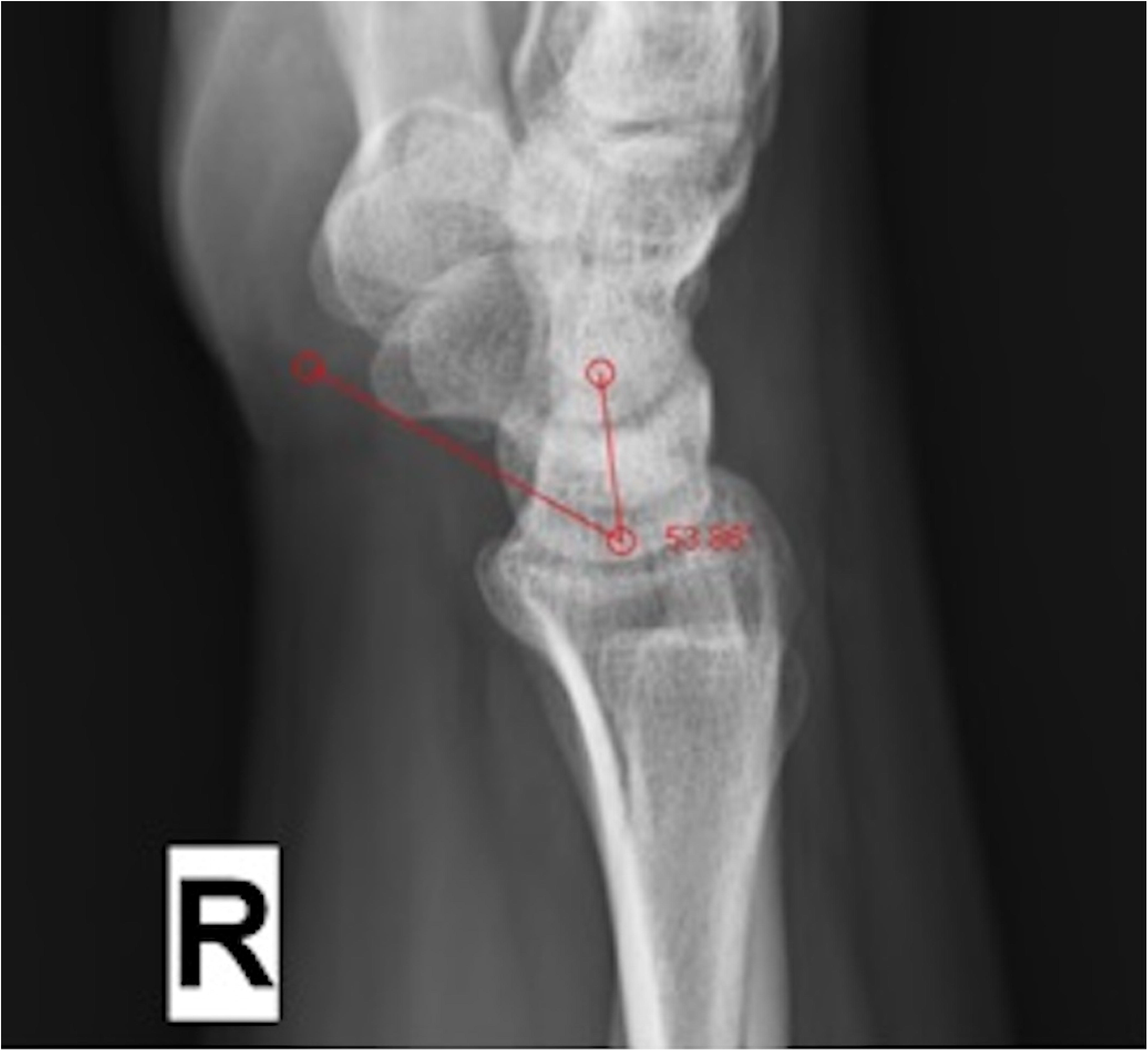
Scapholunate angle subtended between the longitudinal lunate axis and the axis of the scaphoid attained by the tangential method.

## RESULTS

The mean SLA of the 202 lateral radiographs was 51.3° with a standard deviation of 8.87°, ranging between 29° to 74° degrees. The average SLA of female was 51.8° and males 50.8° (table 1). There was no significant difference between the two as indicated by the p-value. There was an increasing trend in the average SLA with age (Table 2).

**Table 1:**
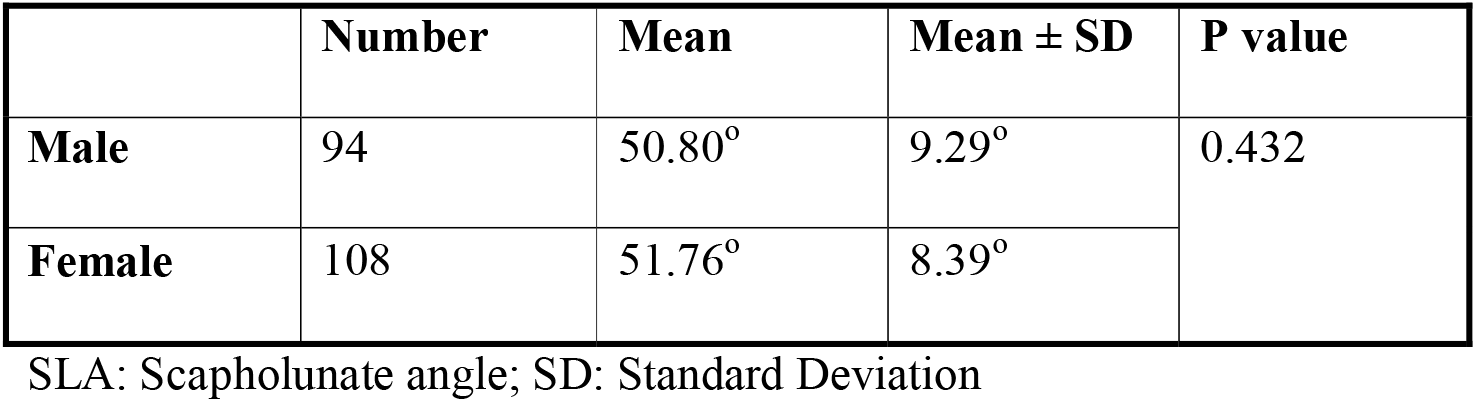
SLA Comparison between Males and Females

**Table 2:** SLA Distribution in different Sub-groups of Age

| Age | Number | Mean | SD | Standard<br>error | Range |  |
| --- | --- | --- | --- | --- | --- | --- |
|  |  |  |  |  | Min | Max |
| **20-30** | 78 | 49.25° | 8.85° | 1.00 | 29.22 | 65.91 |
| **31-40** | 54 | 51.21° | 9.42° | 1.28 | 31.81 | 74.43 |
| **41-50** | 42 | 53.23° | 8.07° | 1.24 | 31.58 | 66.89 |
| **≥ 51** | 28 | 54.52° | 7.29° | 1.37 | 41.05 | 68.25 |
| **Total** | 202 | 51.33° | 8.81° | 0.62 | 29.22 | 74.43 |

SLA: Scapholunate angle; SD: Standard Deviation; Min: Minimum; Max: Maximum

On further investigation it was found that women in the 20-30 years sub-group had a statistically significantly lower mean SLA when compared to the 40-50 years and above 50 years sub-group (table 3). The SLA between the intermediary age groups in the females showed a gradual increase but this was not statistically significant (Fig 3). In case of men, there was no statistical correlation between SLA and age.

**Table 3:**
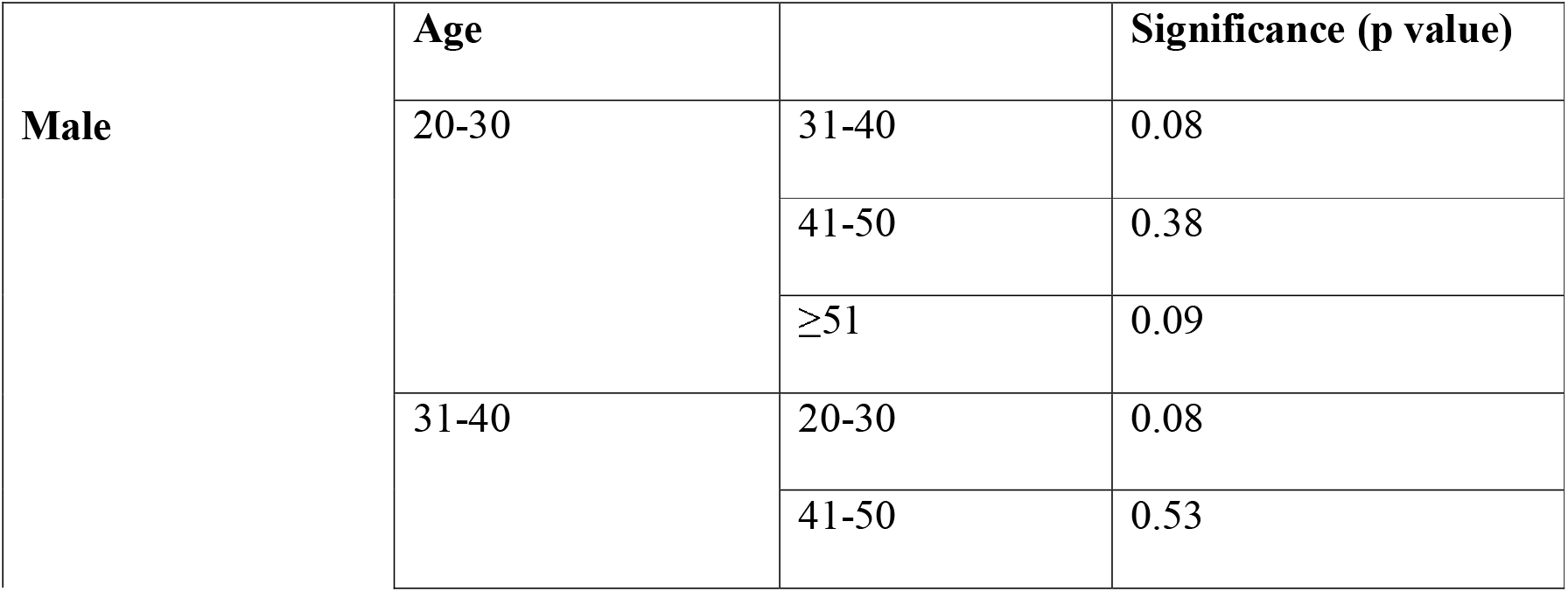
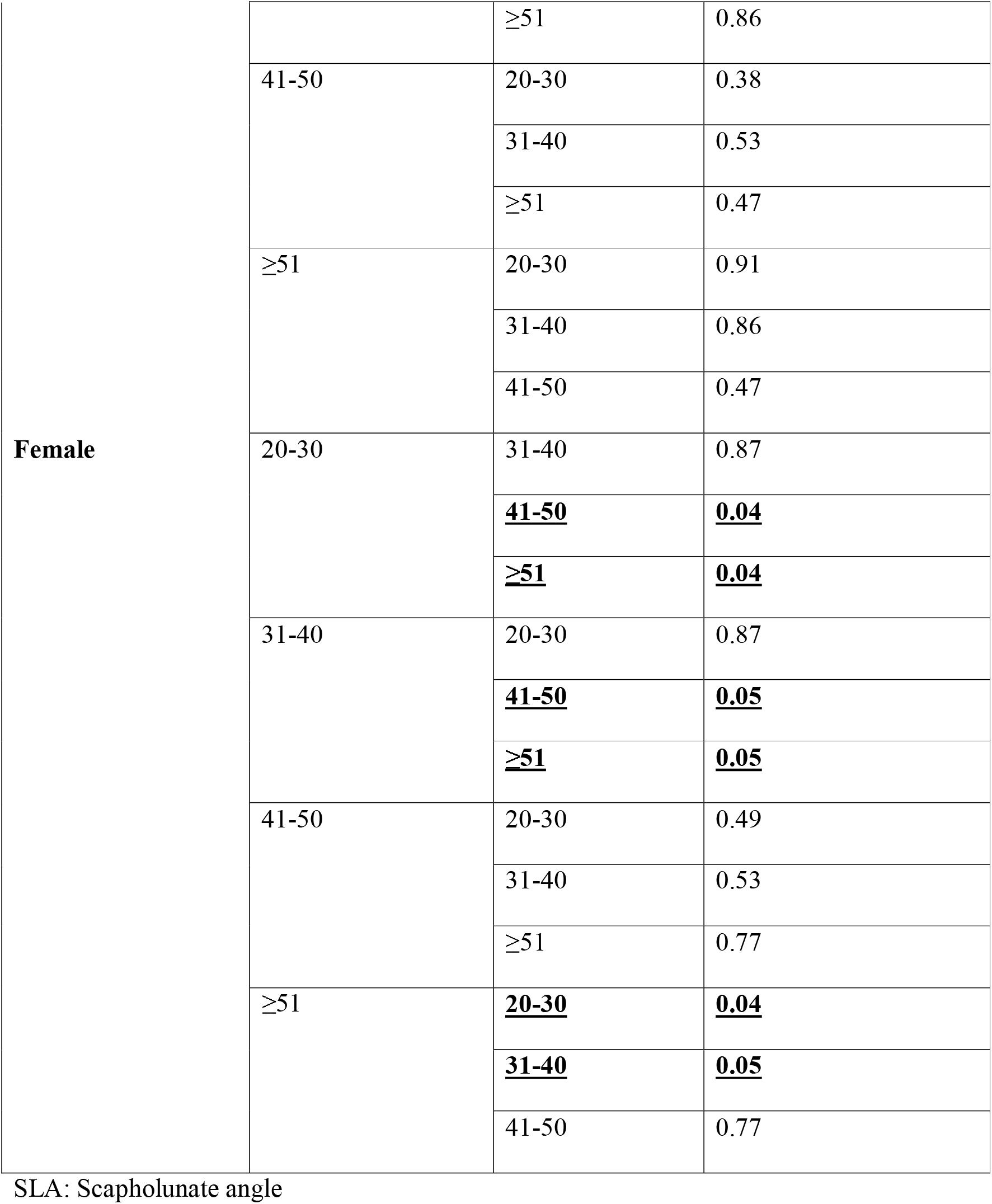
Significance of SLA to Increasing Age in Males and Females

**Fig 3:**
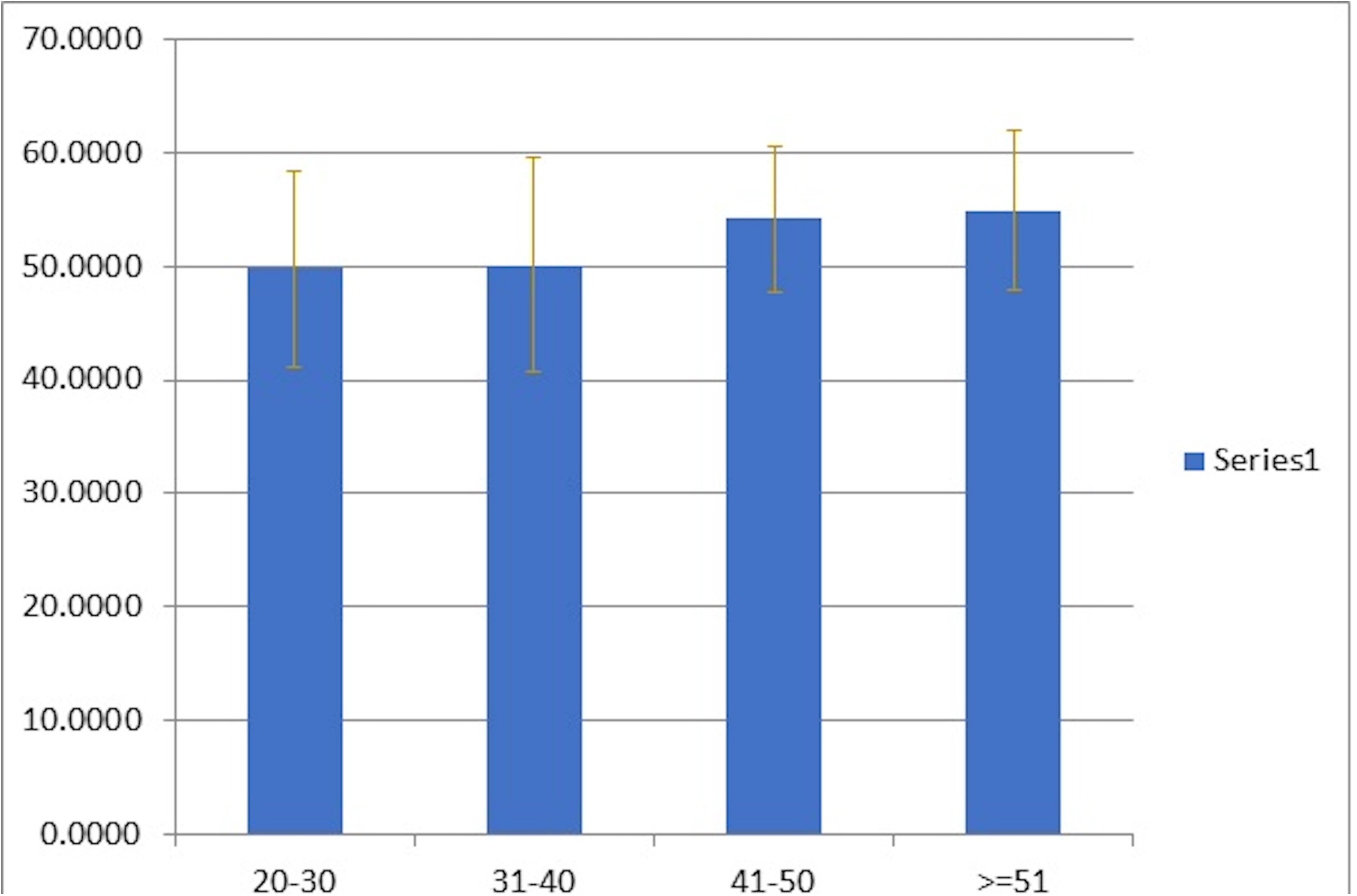
Mean SLA of females in different age group

### Variations among sides and dominance

101 sets of right and left sided wrist radiographs were compared without taking gender or handedness into consideration. When SLA was compared, gender difference was not statistically significant (table 4). Neither was handedness (table 5).

**Table 4:** Difference of SLA in Right and Left side

| | Number | Mean | Mean ± SD | P value |
| --- | --- | --- | --- | --- |
| **Right** | 101 | 50.81° | 8.88° | P = 0.245 |
| **Left** | 101 | 51.85° | 8.76° |  |

SLA: Scapholunate angle; SD: Standard Deviation

**Table 5:** Difference in SLA with Handedness

| | Number | Mean | Mean ± SD | P value |
| --- | --- | --- | --- | --- |
| **Dominant hand** | 101 | 50.73° | 8.86° | P = 0.180 |
| **Non – Dominant hand** | 101 | 51.93° | 8.76° |  |

SLA: Scapholunate angle; SD: Standard Deviation

## DISCUSSION

The scapholunate angle reflects the conditions of scapholunate junction and is used to evaluate carpal bone alignment on lateral radiographs.^12^ On comparing the data of this study with the other international literature available, the mean and range of SLA was in concurrence with study by Sarrafin et al, Larsen et al, Jafari et al. But the range was much narrower in the data by Linsheid et al, Schuid et al and Tang et al when compared to our study (Table 6).

**Table 6:** Comparison of SLA among Different Studies

|  | SLA<br>Average | SLA Range |
| --- | --- | --- |
| S.Indian | 51.3° | 29°-74° |
| Linscheid et al<sup>12</sup> | 46° | 30°-60° |
| Sarrafian et al<sup>12</sup> | 51° | 28°-101° |
| Larsen et al<sup>9, 13</sup> | 50° | - |
| Schuind et al<sup>14</sup> | 45.8°(M) | 40.1°-51.5° |
|  | 48.4°(F) | 40.3°-56.5° |
| Jafari et al, Iran<sup>15</sup> | 51.8° | 30°-80° |
| Tang et al, China<sup>16</sup> | 54.7° | 48.2°-61.2° |

SLA: Scapholunate angle; S: South

There was no significant difference in the average SLA between males and females in any of these studies (Table 7) but the range was much narrower in the data published by Schuhl et al. Again, there was no significant difference in the SLA based on the handedness as compared with the other articles (Table8).

**Table 7:** Comparison of SLA in Genders among the Different Studies

| | Male (Mean ± SD) | Female (Mean ± SD) |
| --- | --- | --- |
| S.Indian (This study) | 50.8° ± 9.29° | 51.76° ± 8.39° |
| Schuind et al<sup>14</sup> | 45.8° ± 5.7° | 48.4° ± 8.1° |
| Schuhl et al<sup>10</sup> | 25.52°-43.32° | 29.2°-49.9° |
| Jafari et al, Iran<sup>15</sup> | 52.4° ± 8.5° | 51.5° ± 8.4° |
| Tang et al, China<sup>16</sup> | There was no significant difference in the SL angle (p = 0.48), |  |

SLA: Scapholunate angle; SD: Standard Deviation; S: South

**Table 8:**
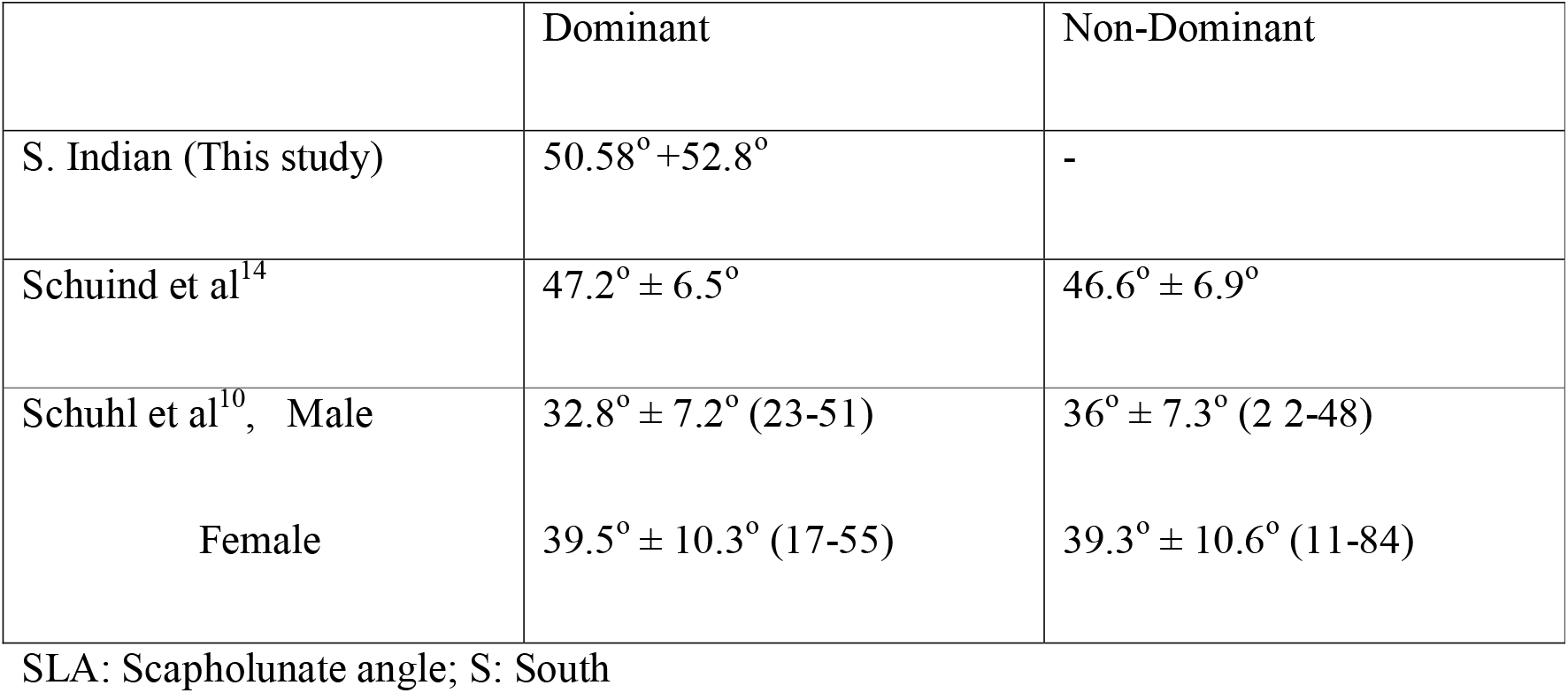
Comparison of SLA with Handedness

On comparing the SLA with age, there was no significance association in the study by Schuind et al^13^ and Jafari et al, Iran^14^ as with the present study (table 9). But an incidental finding in this study was that in case of females, the older women(>40years) had a significantly wider SLA than younger women(20-30years). This was not the same case with men. Another interestingly finding in this study was that 39 out of the 202 (19%) radiographs had SLA more than 60^0^ which is considered as the normal upper limit. It ranged from 61^0^ to 74^0^. All of the 39 subjects were asymptomatic and none of them had an increased scapholunate gap. These interesting observations need further research and evaluation. It is beyond the scope of this paper to explore the reason behind it, but it definitely sparks a question of possibility if it is a common occurrence in many populations which has been missed so long.

**Table 9:** Comparison of SLA with Age

|  | Younger Age | Older Age | Significance |
| --- | --- | --- | --- |
| S.Indian (This study) | 49.25° ± 8.85° | 54.52° ± 7.29° | 0.006 |
| Schuind et al<sup>14</sup> | 50.5° ± 6.2° | 43.7° ± 6.2° | 0.001 |
| Jafari et al, Iran<sup>15</sup> | 51.1° ± 8.8° (20-40yrs) | 52.4° ± 8.0° (40-60 yrs) | NS |
| Tang et al, China<sup>16</sup> | SL angle did not change with patient's age |  |  |

SLA: Scapholunate angle; S: South; NS; Not significant

### Limitation of the study

Further research on SLA is needed in many parts of the world to establish a wide consensus. The follow-up of the individuals especially those with a higher SLA might prove useful, to observe if they are more prone to Scaphotrapeziotrapizoidal (STT) arthritis in the later part of their lives. The significant increase of the SLA in female with age might also be a normal occurrence which has been unnoticed. But again, this needs to be addressed and confirmed with a multicentric study.

## Conclusion

This research paper serves as a pilot study and as a reference for further studies on SL angle in Indians. Though it is in concurrence with the previous articles on SLA, pertaining to the range, average and relation to age sex and handedness, it points out that further research and multicentric studies might be needed to redefine the normal range of SLA. The association of higher SLA with STT arthritis and the significant increase in SLA with age in women require long term cohort studies. The study might also influence designing of implants of carpal bones in the future.

## Data Availability

All data produced in the present study are available upon reasonable request to the authors

## Acknowledgements

Dr.T.Gayathri, Senior Lecturer in Statistics, Department of Allied Health Sciences, Sri Ramachandra Institute of higher Education and Research, Chennai, India

## Funding

This research did not receive any specific grant from funding agencies in the public, commercial or not-for-profit sectors

Written informed consent was obtained from all the volunteers of this study. None of their identities were revealed in the study

## Institutional Ethical Commiittee Approval

This study was approved by the Institutional Ethics Committee (CSP-MED /21/SEP/71/129).

CRediT author statement

Dr.Mohamed Nazir ASHIK: Software, Investigation, Resources, Data Curation, Project administration

Dr.Maithreyi SETHU : Methodology, Software, Validation, Formal analysis, Data Curation, Writing – Original, Writing - Review & Editing

Dr.Srinivasan RAJAPPA : Conceptualization, Resources, Visualization, Supervision

## References

1. Linscheid RL, Dobyns JH, Beabout JW, Bryan RS. Traumatic Instability of the Wrist. The Journal of Bone & Joint Surgery. 1972;54(8):1612–1632.

2. Goldfarb CA, Yin Y, Gilula LA, Fisher AJ, Boyer MI. Wrist Fractures: What the Clinician Wants to Know. Radiology. 2001;219:11–28.

3. Gilula LA, Weeks PM. Post-traumatic Ligamentous Instabilities of the Wrist 1. Radiology. 1978;129(3):641–651.

4. Smith D, Gilula louis, Amadio P. Dorsal Lunate Tilt (DISI Configuration): Sign of Scaphold Fracture Displacement’. Radiology. 1990;176(2):497–499.

5. Mohammed Ali MH. A normal data-base of posteroanterior radiographic measurements of the wrist in healthy Egyptians. Surgical and Radiologic Anatomy. 2009;31(9):665–674. doi:10.1007/s00276-009-0500-4

6. Franklin D, Freedman L, Milne N. Sexual dimorphism and discriminant function sexing in indigenous South African crania. HOMO-Journal of Comparative Human Biology. 2005;55(3):213–228. doi:10.1016/j.jchb.2004.08.001

7. Angermann P, Lohmann M. INJURIES TO THE HAND AND WRIST. A STUDY OF 50,272 INJURIES.

8. Koh KH, Lee HI, Lim KS, Seo JS, Park MJ. Effect of wrist position on the measurement of carpal indices on the lateral radiograph. Journal of Hand Surgery: European Volume. 2013;38(5):530–541. doi:10.1177/1753193412468543

9. Larsen CF, Stigsby B, Mathiesen FK, Lindequist S. Radiography of the wrist: A new device for standardized radiographs. Acta Radiologica. 1990;31(5):459–462. doi:10.1080/02841859009173073

10. Schuhl J, Leroy B, Comtet J, -- CJ. I TUDE RAD.I.OLOGIQUE DE LA MOBILITI RELATIVE DU SCAPHOIDE ET DU SEMI-LUNAIRE RADIOLOGICAL STUDY OF THE MOBILITY OF THE SCAPHO-LUNATE JOINT. Vol 4.; 1985.

11. Capo JT, Accousti K, Jacob G, Tan V. The effect of rotational malalignment on X-rays of the wrist. Journal of Hand Surgery: European Volume. 2009;34(2):166–172. doi:10.1177/1753193408090393

12. Nakamura R, Hori M, Imamura T, Horii E, Miura T. Method for Measurement and Evaluation of Carpal Bone Angles.; 1989.

13. Schuind F, Alemzadeh S, Stallenberg B, Burny F. Does the Normal Contralateral Wrist Provide the Best Reference for X-Ray Film Measurements of the Pathologic Wrist?

14. Jafari D, Taheri H, Shariatzade H, Najd Mazhar F, Jalili A, Ghahramani M. Radiographic Indices in One Hundred Fifty Normal Iranian Wrists. Vol 26.; 2012.

